# Calibration and refinement of ACMG/AMP criteria for variant classification with BayesQuantify

**DOI:** 10.1101/2024.09.08.24313284

**Authors:** Sihan Liu, Xiaoshu Feng, Fengxiao Bu

## Abstract

**Background:** Improving the precision and accuracy of variant classification in clinical genetic testing requires further specification and stratification of the American College of Medical Genetics and Genomics/Association for Molecular Pathology (ACMG/AMP) criteria. While the Clinical Genome Resource (ClinGen) Bayesian framework enables quantitative evidence calibration for selected criteria, standardized tools to optimize evidence thresholds and systematically refine ACMG/AMP criteria remain underdeveloped.

**Methods:** To address this gap, we developed *BayesQuantify*, an R package that provides a unified resource for quantifying evidence strength for ACMG/AMP criteria based on the Bayesian framework. *BayesQuantify* accepts a variant classification file as input and automatically calculates the odds of pathogenicity for each evidence strength, incorporating user-provided prior probabilities of pathogenicity. Through bootstrapping, *BayesQuantify* generates thresholds by aligning the 95% lower boundary of positive likelihood ratio/local positive likelihood ratio values with the odds of pathogenicity for different evidence levels. Three independent datasets (the ClinVar 2019 dataset, the ClinGen curated dataset, and the *PTEN* gene dataset) derived from ClinVar, HGMD, and gnomAD were utilized to evaluate the utility of *BayesQuantify*.

**Results:** Validation across three independent datasets demonstrates that *BayesQuantify* delivers objective, consistent refinements for both categorical and continuous ACMG/AMP evidence. Specifically, we replicated the PP3/BP4 thresholds for four computational tools (BayesDel, VEST4, REVEL, and MutPred2) recommended by the ClinGen Sequence Variant Interpretation Working Group using the ClinVar 2019 dataset. Our analysis also indicated that the PM2 criterion should be downgraded from moderate to supporting evidence, aligning with ClinGen recommendations. Importantly, we have established thresholds for supporting, moderate, and strong evidence for in-silico tools using this tool, thereby expanding the application of PP3/BP4 criteria for missense variants in the *PTEN* gene.

**Conclusions:** *BayesQuantify* is an accessible and user-friendly resource that enhances the rigor and reproducibility of ACMG/AMP criteria application. By facilitating evidence-based stratification and threshold optimization, the tool strengthens variant classification workflows, offering immediate value to clinical genetic testing laboratories and research communities. The package is freely available at https://github.com/liusihan/BayesQuantify.

## Background

Interpreting sequence variants in human genes is a pivotal aspect of diagnosing and managing genetic diseases[1, 2]. However, this task presents considerable challenges due to the complexity and diversity of the genomic data and the lack of standardization and consistency across laboratories[3]. To address these issues, the American College of Medical Genetics and Genomics and the Association for Molecular Pathology (ACMG/AMP) have developed a set of guidelines for variant interpretation, comprising 28 criteria which support or refute the pathogenicity or benignity of a variant[4]. These criteria are grouped into five strength levels of evidence (stand-alone, very strong, strong, moderate, and supporting) and are combined according to specific rules to assign a variant into one of five categories: pathogenic (P), likely pathogenic (LP), uncertain significance (US), likely benign (LB), and benign (B). Since their introduction, the ACMG/AMP guidelines have facilitated the genetic diagnosis of suspected inherited disorders and have become widely adopted by clinical laboratories and researchers as the standard for variant interpretation and reporting[5–8].

In 2018, the ClinGen Sequence Variant Interpretation Working Group (ClinGen SVI) proposed a Bayesian framework that transformed the ACMG/AMP guidelines into a probabilistic system[9]. This framework assumes four levels of evidence and exponentially scaled odds of pathogenicity (OP). It assigns a posterior probability of pathogenicity (Post_P) to a variant by multiplying the OP for each level of evidence (O_PVSt_, O_PSt_, O_PM_, O_PSu,_ O_BVSt_, O_BSt_, O_BM_, O_BSu_) with a prior probability of pathogenicity (Prior_P). The resulting posterior probability is then compared with probability thresholds to categorize variants: (1) the pathogenic combinations would have Post_P > 0.99, (2) the Post_P of the likely pathogenic combinations would be 0.90 < Post_P ≤ 0.99, (3) the Post_P of the likely benign combinations would be 0.001 ≤ Post_P < 0.10, and (4) the one benign category would have Post_P < 0.001. By modelling the ACMG/AMP guidelines with a Prior_P of 0.10 and O_PVSt_ set to 350, 15 out of 17 existing ACMG/AMP evidence combinations (except BA1) were found to be mathematically consistent with the overall framework, validating the approach adopted by the ACMG/AMP.

The Bayesian framework presents opportunities for refining evidence and combining rules, as well as supporting efforts to automate components of variant pathogenicity assessments. Researchers have quantified and extended the strengths of evidence criteria such as PS1/PM5, PS3/BS3, and PP3/BP4 according to the Bayesian framework[10–16]. Importantly, thresholds for PS4, PS4_Moderate, and PS4_Supporting have been quantified by aligning positive likelihood ratio (LR^+^) and local positive likelihood ratio (lr^+^) values with theoretical OP for each evidence strength in our prior study[17]. Our study shows that the refinement of PS4 improves the identification of pathogenic variants, supporting that the specification and stratification of evidence is essential to establish an objective, unbiased and consistent variant classification[7, 8, 18]. However, there is a lack of existing software and tools tailored to quantify the strength of evidence and establish appropriate thresholds to refine the ACMG/AMP criteria.

In this study, we develop *BayesQuantify*, an R package that undergoes the ClinGen Bayesian framework for quantifying the strength of evidence for ACMG/AMP criteria. *BayesQuantify* provides functions to estimate OP for each level of evidence strength and the posterior probability of pathogenicity for the ACMG/AMP combining rules. Furthermore, *BayesQuantify* facilitates the calculation of LR^+^ and lr^+^ for various tested cutoffs, aligning the 95% lower boundary of LR^+^/lr^+^ with OP to establish thresholds. Additionally, users can employ *BayesQuantify* to classify variants in accordance with ACMG/AMP combining rules and Bayesian framework, as well as visualize both the data and statistic results[4, 9]. The *BayesQuantify* is available from GitHub at https://github.com/liusihan/BayesQuantify.

## Methods

### BayesQuantify workflow overview

The functions available through *BayesQuantify* were designed to quantify the evidence strength and thresholds following the algorithm proposed by ClinGen SVI. The package requires one essential input: a *data.frame* containing variants classification results, including the variants, ACMG/AMP evidence, variant classification, and tested features (e.g. Table S1 and Table S2). The step-by-step usage of *BayesQuantify* consists of five main steps as follows (Figure 1):

**Figure 1:**
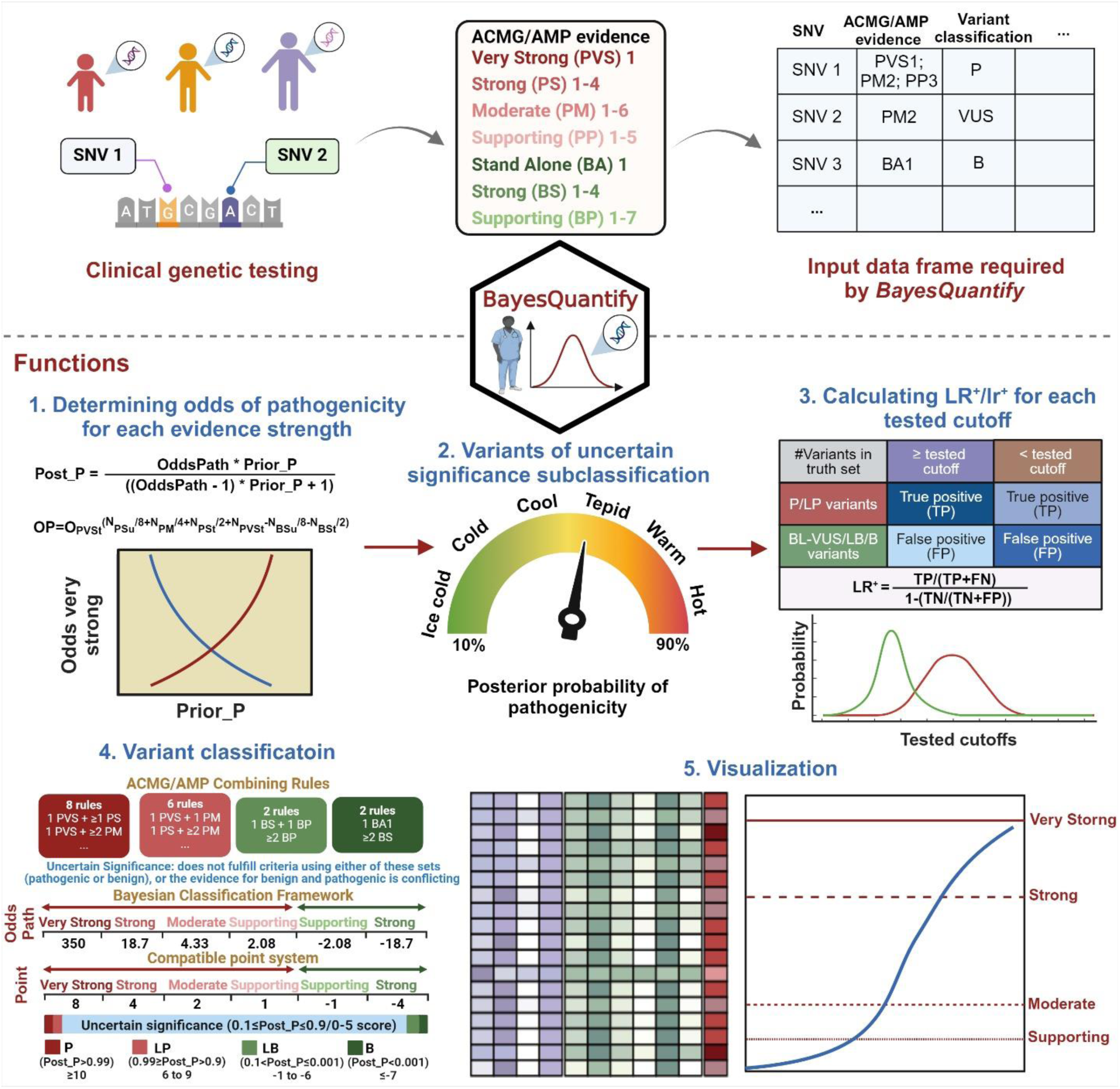
Schematic overview of *BayesQuantify*. *BayesQuantify* begins with the input of variant classification results dataset. Subsequently, it computes the positive likelihood ratio (LR^+^) and local positive likelihood ratio (lr^+^), which are then aligned with predefined odds of pathogenicity (OP) for each evidence strength to refine evidence. Prior_P: prior probability of pathogenicity; Post_P: posterior probability of pathogenicity.

1. **Step 1: Calculation of OP and Post_P for each evidence strength** The SVI Working Group has provided an Excel spreadsheet to calculate Post_P from Prior_P and OP from pathogenic and benign evidence categories[9]. Based on this spreadsheet, users can manually adjust the values of Prior_P and O_PVSt_ to obtain the OP values corresponding to each level of evidence strength. However, this process was computationally inefficient and prone to user bias. To overcome these limitations, we developed *auto_select_postp*(), a core algorithm in *BayesQuantify*, which systematically evaluates O_PVSt_ values over a plausible range (1-10,000) in combination with user-defined Prior_P values. For each iteration, it calculates Post_P using Bayes’ rule and validates compliance with ACMG/AMP rules for combining evidence and predefined pathogenicity probability thresholds. The output identifies the minimum O_PVSt_ value that satisfies these constraints, ensuring robust, reproducible OP and Post_P calculation.

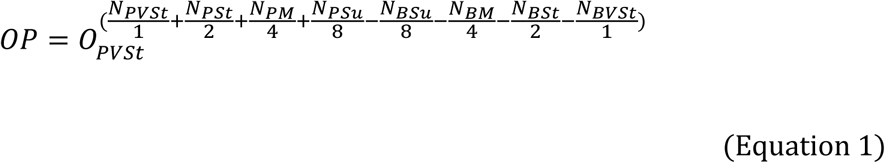

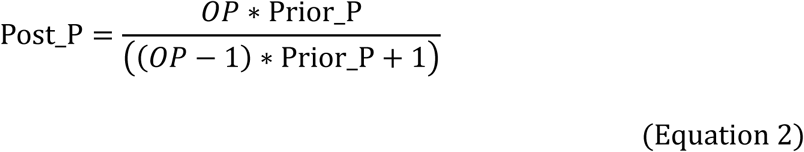
2. **Step 2: Variants of US (VUS) subclassification** The majority of human genomic variants are rare, resulting in a high prevalence of VUS in clinical classifications. To optimize statistical efficiency and reduce classification bias, we have implemented a systematic approach to categorize and filter VUS. The *VUS_classify*() function is developed to stratify variants into six confidence tiers (hot, warm, tepid, cool, cold, ice cold) as defined by the Association for Clinical Genomic Science (ACGS) Best Practice Guidelines (https://www.acgs.uk.com/quality/best-practice-guidelines/#VariantGuidelines). Variants classified as cool, cold, or ice cold are designated benign-leaning VUS (BL-VUS), whereas hot, warm, or tepid variants are classified as pathogenic-leaning VUS (PL-VUS). To establish a robust truth set for refining the evidence criteria, we recommended excluding PL-VUS to avoid confounding the truth set with ambiguous or potentially pathogenic classifications. This stratification ensures a high-confidence dataset for calibrating evidence thresholds while maintaining biological and clinical relevance.
3. **Step 3: Calculating LR^+^/lr^+^** Within the ClinGen Bayesian framework, the OP quantifies the probability of observing specific evidence if a variant is pathogenic. To quantify evidence thresholds, *BayesQuantify* provided functions to calculate the LR^+^/lr^+^ using the following methodologies: For binary variables (e.g. variant marked with PM2 or not): The *LR*() function computes various performance metrics by evaluating the presence or absence of specific evidence among disease-causing and non-pathogenic variants. These metrics include true positive (TP), false positive (FP), true negative (TN), and false negative (FN), overall accuracy, true positive rate (sensitivity), true negative rate (specificity), positive predictive value (PPV), negative predictive value (NPV), F1 score, and LR^+^ (equation 3). The 95% confidence interval (CI) for LR^+^ is estimated using bootstrapping in the R package bootLR [19]. The lower bound of LR^+^ (LR^+^_LB) is then utilized to determine evidence strength by comparing it with the OP thresholds generated in Step 1.

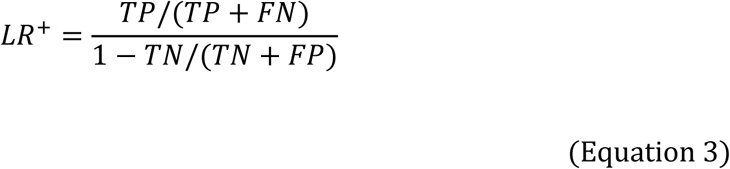 For quantitative variables (e.g. scores from in-silico tools used for tagging PP3/BP4): the *local_bootstrapped_lr()* function first sorts all unique tested cutoffs and positions each cutoff at the center of a user-defined sliding window (default: 0.01). The local posterior probability of pathogenicity and benignity is then calculated for each tested cutoff within the interval, considering a minimum quota of disease-causing and non-pathogenic variants (equation 4). Additionally, the one-sided 95% confidence bound for each estimated lr^+^ is determined through bootstrapping iterations. The *get_lr_threshold()* function can determine the thresholds for each evidence level by mapping the lower bound of local posterior probability with the thresholds generated in Step 1.

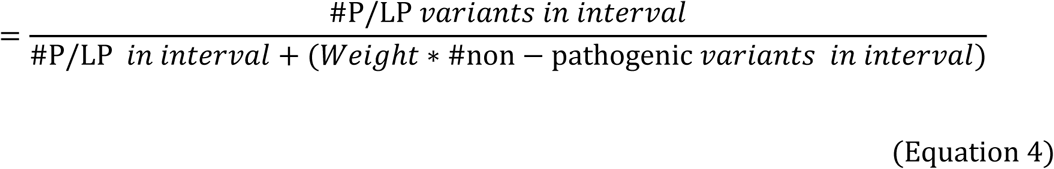
4. **Step 4: Variant classification** *BayesQuantify* offers two functions for variant classification: The *ACMG_Classification*() function categorizes variants into five tiers (P, LP, B, LB, and VUS) using predefined evidence combinations from the ACMG/AMP guidelines; In parallel, the *BCF*() function applies to a Bayesian classification framework to classify variants by calculating posterior probabilities of pathogenicity with user-provided Prior_P and OP for each evidence level.
5. **Step 5: Visualization** To improve user-friendliness, *BayesQuantify* includes several visualization functions: 1) the *multi_plot()* function generates composite plots to display variant characteristics (e.g., allele frequency, conservation scores) in the input data.frame; 2) the *heatmap_LR()* function creates heatmaps to visualize the distribution of positive likelihood ratios across tested cutoffs, highlighting thresholds where LR values meet or exceed OP bounds for pathogenicity or benignity; and 3) the *plot_lr()* function plots local posterior probability curves for quantitative variables, showing the distribution of lr^+^.

All these steps are implemented within the R programming environment to consolidate all intermediate data generated throughout the workflow, ensuring transparency and user-friendly operation. For a detailed explanation of each step with code and examples, see the website of the package (https://github.com/liusihan/BayesQuantify).

### Data sets

#### The ClinVar 2019 dataset

To evaluate the performance of *BayesQuantify*, the ClinVar 2019 dataset, compiled by Pejaver et al. was obtained from https://zenodo.org/records/8347415[10]. This dataset includes missense variants with an allele frequency (AF) below 0.01 in the Genome Aggregation Database (gnomAD v.2.1) and from genes with at least one pathogenic variant of any type in ClinVar[20]. In total, 11,834 variants (5,705 P/LP and 6,129 B/LB variants) from 1,914 genes are included in this dataset (Table S1).

#### The ClinGen Curated Variants dataset

The ClinGen Curated Variants dataset was retrieved from the ClinGen Evidence Repository as of April 7th, 2024 (https://erepo.clinicalgenome.org/evrepo/). This dataset contains expert-curated assertions on variant pathogenicity, along with supporting evidence summaries[18]. It includes classification summaries for 6,768 variants across 74 diseases, with 1,850 P, 1,463 LP, 679 LB, 775 B, and 2,001 US variants (Figure 2 and Table S2). Variants tagged with BA1 were excluded, resulting in the removal of 664 variants from downstream analysis.

**Figure 2:**
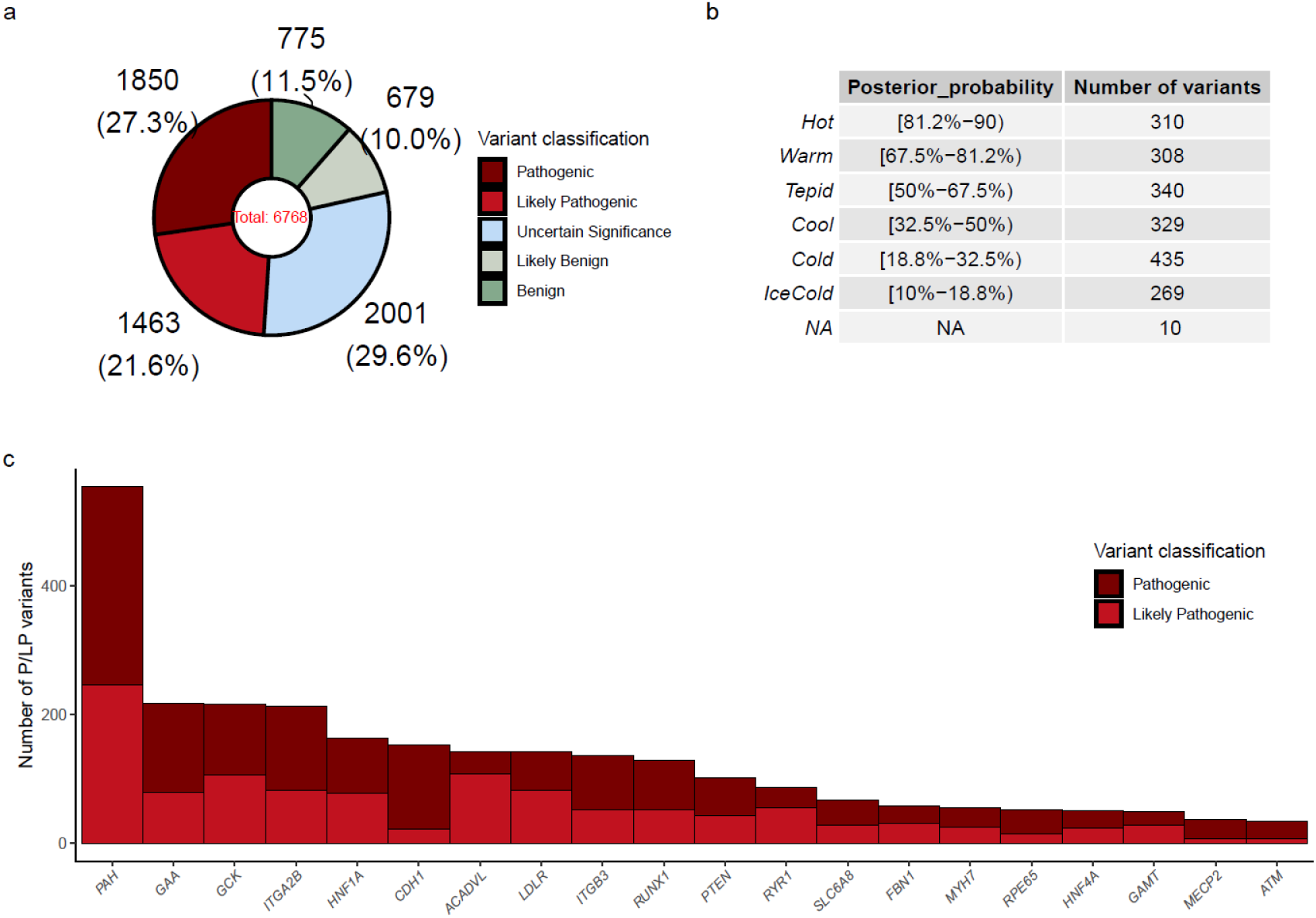
Characteristics of the 6,768 variants in the ClinGen Curated Variants dataset. (a) Summary of classification for the 6,768 variants. (b) Further subclassification of variants of uncertain significance (VUS) to hot, warm, tepid, cool, cold, and ice-cold groups, according to the ACGS Best Practice Guidelines. Variants categorized as “cool/cold/ice cold” were defined as benign-leaning VUS (BL-VUS), while those classified as “hot/warm/tepid” were defined as pathogenic-leaning VUS (PL-VUS). (c) Distribution of pathogenic (P) and likely pathogenic (LP) variants across the top twenty genes.

#### The *PTEN* dataset

To perform the calibration for the *PTEN* gene (MIM# 601728), we constructed a dataset, referred to as the *PTEN* dataset, which included variants classified as disease-causing and non-pathogenic. Variants were sourced from three databases: ClinVar (v20241120)[21], HGMD (2024q1)[22], and gnomAD (v4.1)[20]. These variants were filtered out based on the following criteria: (1) Only missense variants located in the *PTEN* (NM_000314.8) gene with an AF below 0.01 in gnomAD were retained; (2) For variants observed exclusively in gnomAD, only those variants annotated as “PASS” in the “FILTER” column were included; (3) Variants with conflicting pathogenicity annotations between ClinVar (P/LP) and HGMD (DM/DM?) were excluded; and (4) Variants classified as uncertain significance or with conflicting classifications in ClinVar were also removed. Disease-causing variants were defined as P/LP variants, either with a review status of “criteria_provided,_multiple_submitters,_no_conflicts,”, or “reviewed_by_expert_panel,” or those with consistent classifications between ClinVar and HGMD. Non-pathogenic variants were included variants classified as B/LB variants in ClinVar or rare variants only observed in gnomAD (Table S3).

#### Estimation of the prevalence of pathogenic variants for the ClinGen Curated Variants dataset

To estimate the prevalence of pathogenic variants in the ClinGen Curated Variants dataset, we performed the following steps to generate a reference dataset from gnomAD: 1) 332,531 variants were extracted from gnomAD (v4.1), focusing on coding regions of genes with at least one P/LP variant in the ClinGen Curated Variants dataset; 2) variants with an AF < 0.01 and those annotated as “PASS” in the “FILTER” column were retained; and 3) P/LP variants present in ClinVar (v20241120) or HGMD (2024q1) were removed. The final reference dataset consisted of 133,270 potentially non-pathogenic variants. Based on this, we estimated the prevalence of pathogenic variants in the ClinGen Curated Variants dataset to be 2.48% (3,313/133,270).

#### Estimation of the prevalence of pathogenic variants for the *PTEN* dataset

For estimating the prevalence of pathogenic variants in the *PTEN* dataset, we utilized a combined dataset from ClinVar (v20241120), HGMD (2024q1), and gnomAD (v4.1). Variants in the *PTEN* gene with an AF > 0.01 in gnomAD, as well as those with conflicting pathogenicity annotations between ClinVar (P/LP) and HGMD (DM/DM?), were excluded. The prevalence of pathogenic variants in this dataset was found to be 4.68% (1,319/28,171).

#### Calibration of computational tools in the *PTEN* dataset

To systematically evaluate and calibrate computational tools, we annotated variants using precomputed prediction scores from the Database of Human Nonsynonymous SNPs (dbNSFP 4.7a) via the Variant Effect Predictor (VEP) [23, 24]. A total of 38 prediction scores from the dbNSFP database, covering the entire human genome, were compared. All genomic positions for these precomputed scores were based on the GRCh38/hg38 reference genome. To standardize the assessment, we employed a rank score methodology, which scaled the raw prediction scores to a 0-1 range, reflecting the proportion of variants deemed less damaging. The performance of these 38 tools was then evaluated in the *PTEN* dataset using receiver-operating characteristic (ROC) curves, generated with the R package pROC[25].

## Results

### Generating OP values and high-confidence datasets using *BayesQuantify*

Comprehensive OP values for ACMG/AMP criteria were established across three distinct datasets through the *auto_select_postp*() function in *BayesQuantify*. For the ClinVar 2019 dataset (Prior_P=0.0441, estimated by Pejaver et al.), pathogenic evidence required OP values ≥2.41 (Supporting), ≥5.80 (Moderate), ≥33.65 (Strong), and ≥1132 (Very Strong), while benign evidence thresholds were ≤0.415, ≤0.172, ≤0.03, and ≤0.001 respectively. The ClinGen Curated Variants dataset (Prior_P=0.0248) exhibited elevated pathogenic thresholds (O_PSu_≥2.65, O_PM_≥7.06, O_PSt_≥49.9, O_PVSt_≥2490) with corresponding benign thresholds at O_BSu_≤0.376, O_BM_≤0.142, O_BSt_≤0.02, and O_BVSt_≤0.0004. For the *PTEN* dataset, the OP values were: O_PSu_ ≥ 0.384, O_PM_ ≥ 5.682, O_PSt_ ≥ 32.28, O_PVSt_ ≥ 1042, O_BSu_ ≤ 0.42, O_BM_ ≤ 0.176, O_BSt_ ≤ 0.031, and O_BVSt_ ≤ 0.001, with a Prior_P of 0.0468.

To accurately refine the ACMG/AMP criteria, the *VUS_classify()* function was used to categorize variants of uncertain significance into six levels, generating high-confidence datasets by excluding PL-VUS variants. In the ClinGen Curated Variants dataset, this approach resulted in 5,137 variants, comprising 1,847 P, 1,463 LP, 123 B, 673 LB, and 1,031 BL-VUS variants. Correspondingly, the ClinVar 2019 dataset encompassed 5,705 disease-causing variants and 6,129 non-pathogenic variants, while the *PTEN* dataset consisted of 326 disease-causing variants and 69 non-pathogenic variants.

### Validating the utility of *BayesQuantify* for evidence refinement and variant classification

To assess the accuracy of *BayesQuantify* in refining evidence for variant classification, we replicated the calibration of four recommended computational tools (BayesDel, MutPred2, REVEL, and VEST4) using the ClinVar 2019 dataset, as proposed by Pejaver et al. for determining PP3 and BP4 evidence levels. Using 10,000 bootstrap iterations, we calculated local posterior probabilities for each unique score generated by these tools. Our analysis confirmed that all four tools achieved thresholds for strong evidence of pathogenicity (BayesDel ≥ 0.53, MutPred2 ≥ 0.925, REVEL ≥ 0.921, VEST4 ≥ 0.96). For benignity, BayesDel (≤ -0.36) and VEST4 (≤ 0.32) reached moderate evidence thresholds, while MutPred2 (≤ 0.02) and REVEL (≤ 0.04) met strong evidence thresholds (Figure 3, Table S4). These results align with Pejaver et al.’s findings, validating the reliability of *BayesQuantify* in evidence calibration.

**Figure 3:**
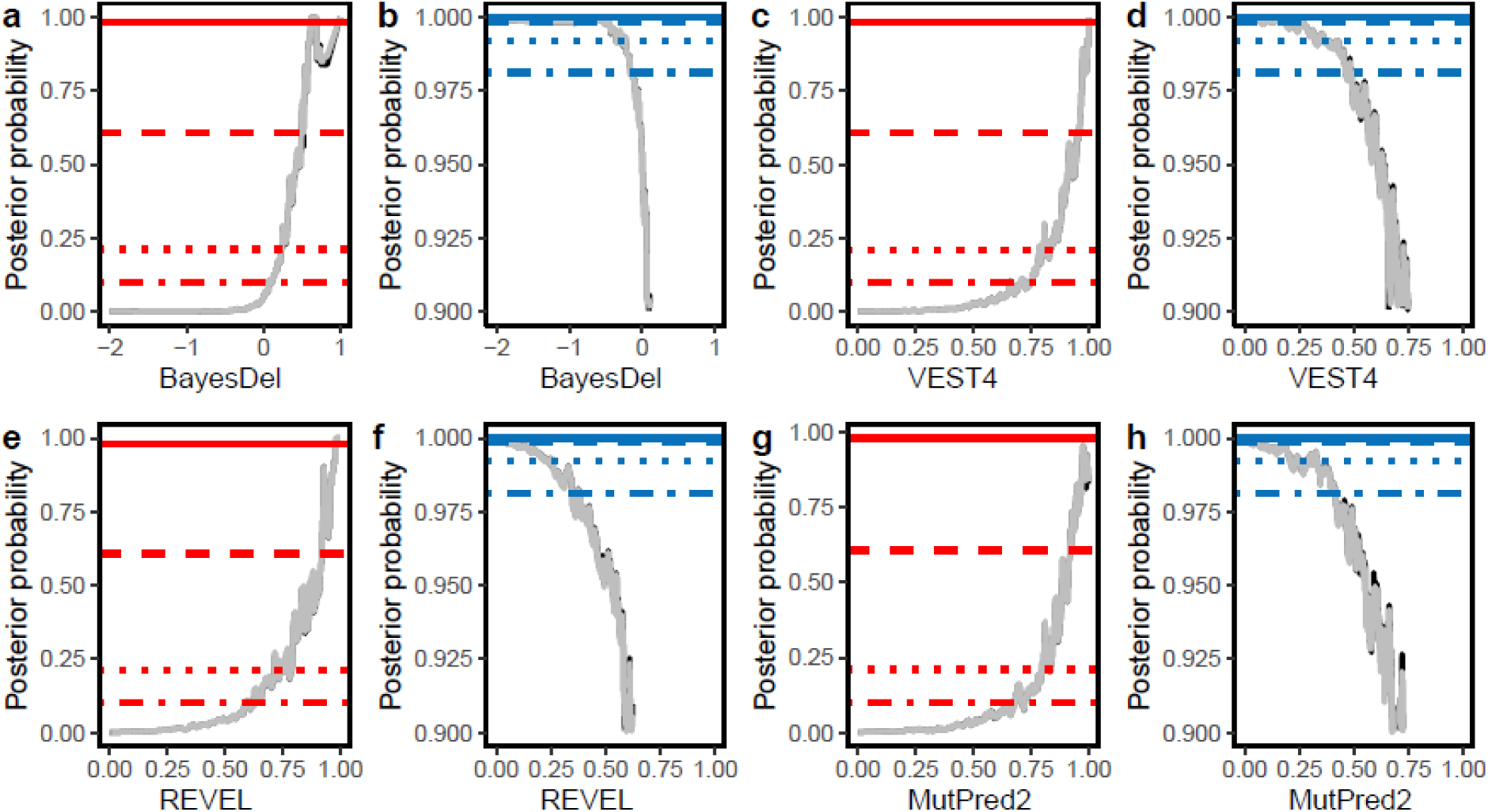
Local posterior probability curves of pathogenicity and benignity for various BayesDel, VEST4, REVEL and MutPred2 scores in the ClinVar 2019 dataset. The horizontal lines depict the posterior probability thresholds for supporting, moderate, strong, and very strong evidence. The black curves represent the posterior probability estimated from ClinVar 2019 dataset, while the grey curves represent one-sided 95% confidence intervals calculated from 10,000 bootstrap samples of this dataset.

We further evaluated *BayesQuantify*’s ability to classify variants into five categories (P, LP, B, LB, and VUS) according to the ACMG/AMP guidelines and the Bayesian classification framework. Using the ClinVar 2019 dataset, *BayesQuantify* demonstrated 99.02% concordance (n = 6,702 variants) with existing classifications (Table S5). Of the 66 discordant cases, discrepancies arose from clinical judgement beyond guideline-based evidence. A representative example is the *CDH23* variant NM_022124.6:c.380A>G (p.Asp127Gly), which is classified as a VUS according to the ACMG/AMP criteria. This variant is absent in gnomAD v2.1.1 (PM2_Supporting) and computationally predicted to be deleterious (REVEL = 0.796, PP3). Although detected in an individual with autosomal recessive Usher syndrome (PP4) and confirmed to be in compound heterozygosity with a pathogenic frameshift variant (NM_022124.6:c.1949dup, p.Leu651fs; PM3), its Bayesian posterior probability (0.812) fell below the threshold for likely pathogenic classification (≥ 0.9). Despite this, the ClinGen Hearing Loss Variant Curation Expert Panel (VCEP) re-evaluated the variant on 18 January 2023 and retained its likely pathogenic designation based on clinical expertise.

### Evaluating the clinical weight of PM2 criteria in variant pathogenicity classification

The 2015 ACMG/AMP guidelines originally defined PM2 as a moderate pathogenic criterion, applicable when a variant is absent or extremely rare (e.g., recessive conditions) in population databases such as gnomAD. However, concerns arose about overestimating its predictive value, as large-scale studies (e.g., ExAC) revealed that 54% of rare variants occurred only once, suggesting that rarity alone is insufficient to infer pathogenicity. To address this concern, we evaluated the clinical weight of PM2 criteria using *BayesQuantify*. In the ClinGen Curated Variants dataset, PM2 was designated as moderate evidence and supporting evidence in accordance with gene-/disease-specific guidelines and emerged as the most frequently applied pathogenic criterion, being utilized in 3,072 P/LP variants, 580 BL-VUS, and 114 B/LB variants, respectively (Figure S1). We evaluated whether or not PM2 and PM2_Supporting met the relative odds of pathogenicity for a Moderate (7.064:1) and Supporting pathogenic (2.658:1) evidence type. Consequently, a total of 3,766 variants were tagged with PM2_Supporting, including 3,072 P/LP variants and 694 BL-VUS/B/LB variants. The accuracy was 0.819, PPV was 0.816, sensitivity was 0.928, specificity was 0.620, and LR^+^ was 1.497 (95% CI, 1.443–1.555), which was lower than the O_PSu_ value for the supporting evidence level (Table 1 and Table S6). Furthermore, 1,102 variants were identified with a moderate evidence level of PM2, comprising 1,002 P/LP variants and 100 BL-VUS/B/LB variants. However, the accuracy was 0.531, PPV was 0.910, sensitivity was 0.302, specificity was 0.945, and LR^+^ was 5.531 (95% CI, 4.575–6.818), reaching the supporting evidence level (LR^+^_LB ≥ 2.658).

**Table 1:** Posterior probability and positive likelihood ratio values corresponding to each evidence strength in the three datasets included in this study.

| **Evidence Strength** | **ClinGen Curated Variants dataset** |  | **ClinVar 2019 dataset** |  | ***PTEN* dataset** |  |
| --- | --- | --- | --- | --- | --- | --- |
|  | **OP** | **Posterior probability** | **OP** | **Posterior probability** | **OP** | **Posterior probability** |
| PVS | 2490 | 0.985 | 1132 | 0.981 | 1042 | 0.981 |
| PS | 49.90 | 0.560 | 33.645 | 0.608 | 32.28 | 0.613 |
| PM | 7.064 | 0.153 | 5.800 | 0.211 | 5.682 | 0.218 |
| PP | 2.658 | 0.064 | 2.408 | 0.100 | 2.384 | 0.105 |
| BP | 0.376 | 0.990 | 0.415 | 0.981 | 0.420 | 0.980 |
| BM | 0.142 | 0.996 | 0.172 | 0.992 | 0.176 | 0.991 |
| BS | 0.020 | 0.999 | 0.030 | 0.999 | 0.031 | 0.998 |
| BVS | 0.0004 | 1.000 | 0.001 | 1.000 | 0.001 | 1.000 |

### Calibration of in-silico tools for predicting pathogenicity for the *PTEN* gene

We systematically evaluated 38 variant impact prediction algorithms curated in the dbNSFP v4.2a database to establish calibrated thresholds for *PTEN* missense variant pathogenicity classification, enhancing precision in clinical interpretation aligned with ACMG/AMP guidelines. The distribution of prediction scores between non-pathogenic and disease-causing variants in the *PTEN* dataset across all 38 tools was shown in Figure S2. Twelve tools (SiPhy, phastCons, phyloP, GERP++, fitCons, GenoCanyon, fathmm_MKL, DANN, LIST_S2, fathmm_XF, MutationTaster, and LRT) were excluded from downstream analysis due to over-mixed prediction scores for both non-pathogenic and disease-causing variants. The remaining 26 tools showed an area under the curve (AUC) values ranging from 0.782 to 0.946 (Figure S3), with the Evolutionary model of Variant Effect (EVE; AUC =0.946) exhibiting the best performance, followed closely by Rare Exome Variant Ensemble Learner (REVEL; AUC = 0.945) and Variant Effect Scoring Tool (VEST; AUC = 0.943).

To assess the clinical utility of these prediction scores for variant curation based on ACMG/AMP recommendations, we calibrated the top 10 performing tools. The local posterior probability-based approach proposed by ClinGen SVI allowed for identifying thresholds corresponding to different levels of evidence. This approach was applied to the following 10 tools: AlphaMissense[26], BayesDel[27], ClinPred[28], EVE[29], M-CAP[30], MetaLR[31], MetaRNN[32], MVP[33], REVEL[34], and SIFT[35]. We obtained variant scores from these tools, calculated local posterior probabilities, and assigned corresponding strengths of evidence based on the thresholds outlined in Table 1.

We identified thresholds for supporting and moderate evidence of pathogenicity (PP3) and benignity (BP4) for all tools, except for MetaLR and MVP, which did not yield moderate evidence for BP4. Interestingly, the local posterior probability curves demonstrated that, at appropriate thresholds, several tools could provide strong evidence for pathogenicity (ClinPred, M-CAP, MetaLR, MetaRNN) or benignity (AlphaMissense), as illustrated in Figure 4 and Table 2.

**Figure 4:**
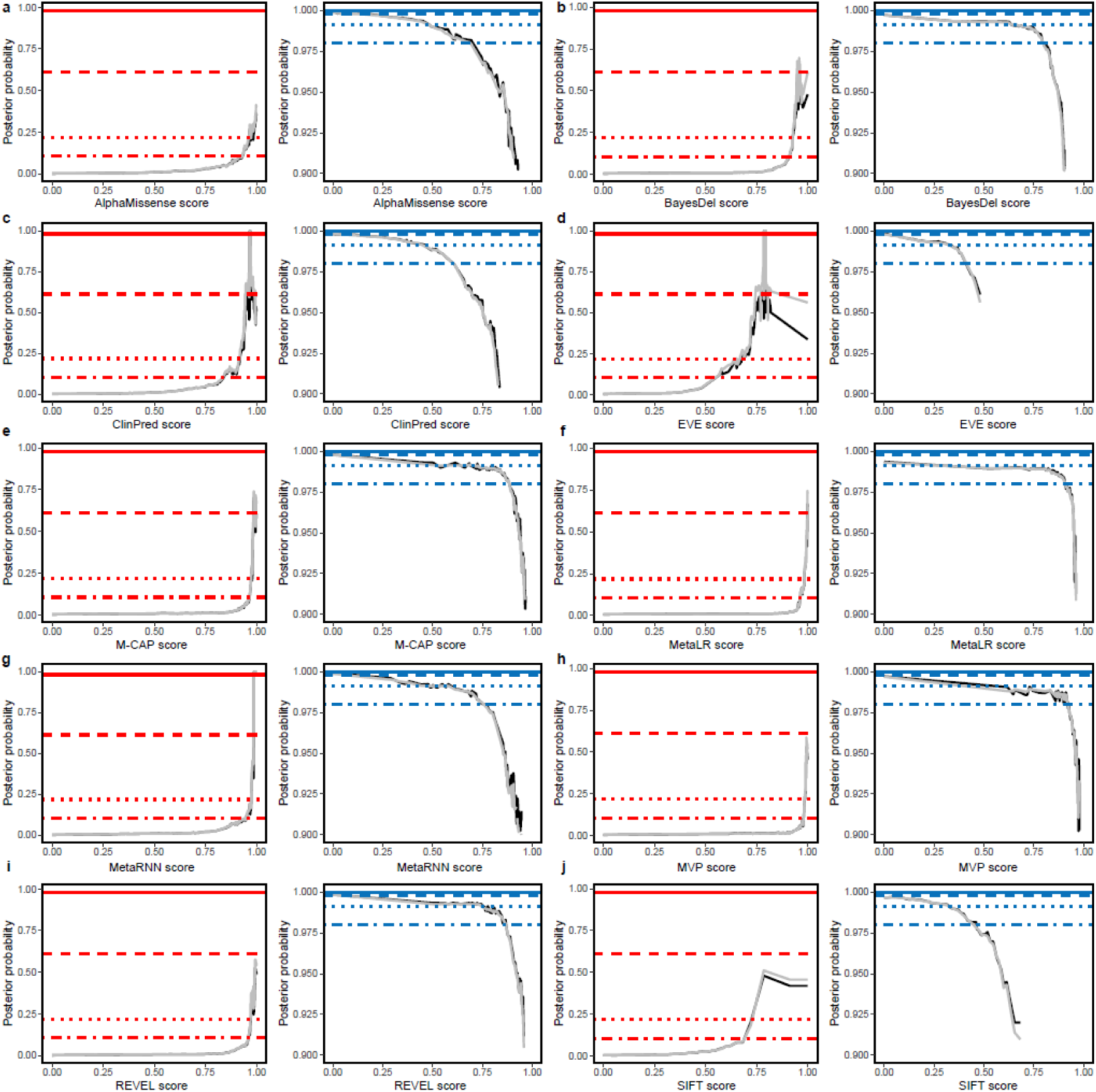
Local posterior probability curves ten in-silico tools in the *PTEN* dataset. The horizontal lines depict the posterior probability thresholds for supporting, moderate, strong, and very strong evidence. The black curves represent the posterior probability estimated from *PTEN* dataset, while the grey curves represent one-sided 95% confidence intervals calculated from 10,000 bootstrap samples of this dataset. The points at which the gray curves intersect the horizontal lines represent the thresholds for the relevant intervals.

**Table 2:** Estimated threshold ranges corresponding to the four pathogenic and four benign intervals for the *PTEN* gene.

| Method | Benign (BP4) |  |  |  | Pathogenic (PP3) |  |  |  |
| --- | --- | --- | --- | --- | --- | --- | --- | --- |
|  | Very Strong | Strong | Moderate | Supporting | Supporting | Moderate | Strong | Very Strong |
| AlphaMisse<br>nse | - | $\leq 0.060$ | (0.060, 0.453] | (0.453, 0.643] | [0.935, 0.974) | $\geq 0.974$ | - | - |
| BayesDel | - | - | $\leq 0.638$ | (0.638, 0.794] | [0.908, 0.925) | $\geq 0.925$ | - | - |
| ClinPred | - | - | $\leq 0.447$ | (0.447, 0.564] | [0.836, 0.917) | [0.917, 0.999) | $\geq 0.999$ | - |
| EVE | - | - | $\leq 0.324$ | (0.324, 0.385] | [0.567, 0.668) | $\geq 0.668$ | - | - |
| M-CAP | - | - | $\leq 0.686$ | (0.686, 0.883] | [0.967, 0.973) | [0.973, 0.998) | $\geq 0.998$ | - |
| MetaLR | - | - | - | $\leq 0.907$ | [0.965, 0.983) | [0.983, 0.999) | $\geq 0.999$ | - |
| MetaRNN | - | - | $\leq 0.581$ | (0.581, 0.745] | [0.949, 0.976) | [0.976, 0.986) | [0.986, 0.987) | $\geq 0.987$ |
| MVP | - | - | - | $\leq 0.912$ | [0.980, 0.985) | $\geq 0.985$ | - | - |
| REVEL | - | - | $\leq 0.744$ | (0.744, 0.865] | [0.959, 0.972) | $\geq 0.972$ | - | - |
| SIFT | - | - | $\leq 0.328$ | (0.328, 0.440] | [0.722, 0.785) | $\geq 0.785$ | - | - |

## Discussion

The ClinGen Bayesian framework offers a solid mathematical basis for refining ACMG/AMP criteria within clinical laboratory practice. However, to our knowledge, there is a lack of freely available software tools that provide similar functionalities for the academic community. In this article, we present *BayesQuantify*, an R package designed to enhance the flexibility and convenience of quantifying the strength of evidence and establishing corresponding thresholds for ACMG/AMP criteria based on the Bayesian framework. It is important to note that while *BayesQuantify* is a computational tool, it necessitates human input to ensure the accuracy of its results. By expanding the evidence base of the ACMG/AMP criteria, *BayesQuantify* has the potential to elevate the classification of VUS to definitive interpretations.

*BayesQuantify* offers an objective assessment of whether various forms of evidence align with the relative weights specified by the Bayesian framework. Our analysis indicated that the PM2 criterion should be downgraded from moderate to supporting evidence, which is consistent with the ClinGen SVI recommendations (https://clinicalgenome.org/docs/pm2-recommendation-for-absence-rarity/). This adjustment addresses the historical overestimation of the predictive value of PM2 as population databases reveal high rates of ultra-rare variants. To mitigate the potential impact of PM2 downgrading in variant classification, the ClinGen SVI also proposes novel evidence combinations beyond the 2015 ACMG/AMP guidelines, combining a Very Strong (e.g. PVS1) and a Supporting criterion (e.g. PM2_Supporting) to achieve a Likely Pathogenic classification (Bayesian posterior probability=0.988). This approach ensures that novel loss-of-function variants with strong mechanistic evidence (PVS1) and an adjusted PM2 weight retain actionable classifications, while mitigating over-reliance on rarity alone.

The Bayesian framework implemented in *BayesQuantify* extends the quantification of evidence strength from categorical to continuous variables, including computational predictors. We corroborates Pejaver et al.’s findings, demonstrating that BayesDel, MutPred2, REVEL and VEST4 achieve strong evidence thresholds for pathogenicity (PP3) and moderate evidence for benignity (BP4). The slight discrepancies between our results and those of Pejaver et al. can be attributed to variations in the bootstrap process. Although these findings are applicable across various genes, a genome-wide approach may mask the heterogeneous performance of predictors within distinct genes[36, 37]. As such, gene-specific calibrations are crucial for improving accuracy and precision in variant classification.

Our study attempted to calibrate the PP3 and BP4 criteria for missense variants in the *PTEN* gene, thereby refining their application in variant classification. Germline pathogenic variants in *PTEN* are associated with a highly heterogeneous clinical spectrum and are observed in individuals with conditions such as Cowden syndrome, Bannayan-Riley-Ruvalcaba syndrome, and other phenotypes, including macrocephaly and autism, collectively referred to as PTEN Hamartoma Tumor Syndrome[38, 39]. In 2018, the ClinGen PTEN Expert Panel introduced gene-specific criteria for *PTEN* variant curation, incorporating disease-specific modifications and adjustments to the strength of evidence across 19 criteria[40]. However, the guidelines did not establish a set of thresholds for applying PP3 or BP4 criteria to *PTEN* missense variants, leaving a critical gap in variant interpretation. According to our results, we recommend using MetaRNN or AlphaMissense to determine evidence for codes PP3 and BP4, as these tools provide strong or moderate evidence for pathogenicity and benignity[26, 32]. Both tools demonstrated high AUC values (above 0.93), maximizing the strength of evidence while minimizing false positives in the supporting and moderate categories. Given the absence of a dedicated validation dataset, future studies are necessary to validate these thresholds.

A limitation worth discussing is the assumption within the Bayesian framework that each categorical type of evidence at the same evidence strength level had the same mathematical support. Given the complexity of interpreting genetic evidence, this simplifying assumption is reasonable but may not fully capture the nuances involved. Previous studies have identified dependencies and correlations between evidence rules, such as those observed in BS3/BP4 and PVS1/PP3[41, 42]. To address this limitation, extending the Bayesian framework by estimating more precise evidence weights using real-world datasets containing true positives and true negatives could provide greater accuracy. Such methods have already been implemented for some evidence types in genes related to breast and colorectal cancers[16, 43]. Furthermore, given the potential misclassification in public databases[44–46], we emphasize the importance of user-curated datasets in refining evidence when utilizing *BayesQuantify*.

## Conclusions

In summary, we have developed *BayesQuantify*, an R package for calibrating and refining ACMG/AMP criteria for variant classification according to the Bayesian framework. This tool is user-friendly and significantly facilitates the shift from a predominantly subjective assessment to a more quantitative and objective approach. By expanding the evidence base of the ACMG/AMP criteria, *BayesQuantify* can enhance the existing ACMG criteria and improve the reclassification of VUSs.

## Data Availability

The ClinGen Curated Variants data was downloaded from https://erepo.clinicalgenome.org/evrepo/. The ClinVar 2019 set was sourced from https://zenodo.org/records/8347415. The three datasets included in this study are provided in the supplementary tables and are also included in the BayesQuantify package.

## Abbreviations

ACMG/AMP: American College of Medical Genetics and Genomics/ Association for Molecular Pathology
AF: Allele frequency
B: Benign
BL-VUS: Benign-leaning VUS
CI: Confidence interval
gnomAD: The Genome Aggregation Database
LB: Likely Benign
LP: Likely Pathogenic
LR^+^: Positive likelihood ratio
lr^+^: Local positive likelihood ratio
OP: Odds of pathogenicity
O_PVSt_: Odds of pathogenicity for “very strong” evidence
P: Pathogenic
Post_P: Posterior probability of pathogenicity
Prior_P: Prior probability of pathogenicity
VUS: Variants with Uncertain Significance

## Declarations

### Ethics approval and consent to participate

Not applicable

### Consent for publication

Not applicable

### Availability of data and materials

The ClinGen Curated Variants data was downloaded from https://erepo.clinicalgenome.org/evrepo/. The ClinVar 2019 set was sourced from https://zenodo.org/records/8347415. The three datasets included in this study are provided in the supplementary tables and are also included in the *BayesQuantify* package.

### Authors’ contributions

**Sihan Liu**: Conceptualization, Methodology, Validation, Investigation, Visualization, Writing – original draft, Writing – review & editing, Supervision. **Xiaoshu Feng**: Methodology, Writing – original draft, Writing – review & editing. **Fengxiao Bu**: Supervision, Writing – original draft, Writing – review & editing. All authors have read and approved the final manuscript.

## Acknowledgements

We would like to extend a special thanks to Information Center of West China Hospital for technical support. Figure 1 created with BioRender.com.

